# Toll-like Receptor 9 in breast carcinoma is a good prognostic marker in patients treated with neoadjuvant chemotherapy

**DOI:** 10.1101/2020.05.28.20114850

**Authors:** Aradhana Singh, Arghya Bandyopadhyay, Narendranath Mukherjee, Anupam Basu

## Abstract

**Purpose:** TLR9 is the sensor of fragmented nucleic acid signature as a part of innate immune surveillance. TLR9 can recognize the DNA fragments released from the chemotherapy-treated cancer cells in tumour tissue and induce an inflammatory response.

The aim of this was toinvestigate the prognostic importance and survivability benefit of TLR9 expression in breast cancer patients treated with neoadjuvant chemotherapy.

**Methods:** Expression of TLR9 in breast carcinoma samples was studied in two patient cohorts, with neoadjuvant chemotherapy (NACT), and without NACT, by immunohistochemistry. Expression of TLR9 was analysed in relation to prognosis, overall survivability as well as risk factor analysis for neoadjuvant chemotherapy treatment using web-tools like SurvExpress and K-M Plotter.

**Results:** TLR9 was expressed in malignant epithelial cancer cells as well as in adjacent stromal cells. TLR9 in malignant epithelial cells was significantly high in patients treated with neoadjuvant chemotherapy compared to the patients without neoadjuvant chemotherapy. The prognostic and survival analysis by SurvExpress and Kaplan-Meier plotter demonstrated that high TLR9 expression is related to better overall survival in patients treated with NACT.

**Conclusions:** Thus, we are showing for the first time that TLR9 is good prognostic marker in breast cancer treated with neoadjuvant chemotherapy and can be used for the selection of the neo-adjuvant regime.

## 1 Introduction

Toll-like receptor 9 (TLR9) is an endosomal innate immune system receptor for unmethylated bacterial or viral CpG motif-containing DNA[1]. The cellular location of TLR9 plays a decisive role in discriminating the identity of self and non-self-nucleic acid. Stimulation of TLR 9 induces an NF-κB mediated rapid inflammation with the expression of various interleukins and cytokines that eventually activates the adaptive immune system and clearance of invading pathogen [2],[3]. A similar inflammatory response mediated via TLR9 also occurs in tissue damage such as infarction or trauma [4,5]. TLR9 is constitutively expressed in B-cells and in the type I INF-producing plasmacytoid dendritic cells (pDCs), but upon activation, additional immune cells like human neutrophils, monocytes, and monocyte-derived CD4+ T-cells have also been shown to express the receptor [6,7,8,9]. In a recent study, Tuomela et al., (2013) [10] demonstrated tissue-derived DNA fragments released from the chemotherapy-treated dead cancer cells are taken up by surviving cells of the same tumor, in which TLR 9 –mediated recognition of the DNA results in the inflammatory response. The expression of TLR 9 in breast tumor tissues has also been reported earlier [11]. In locally advanced breast cancer (LABC), neoadjuvant chemotherapy (NACT) is increasingly used to induce tumor shrinkage that allows smaller surgical resection. Neo-adjuvant administration of FEC (Fluorouracil, epirubicin, and cyclophosphamide) followed by paclitaxel and/or trastuzumab partly exert their antitumor activity by induction of an antitumor immune response aimed at cells injured by chemotherapy [12,13]. Neo-adjuvant administration therapy is a better treatment response that has been shown in different breast cancer studies [14,15]. It has also been shown in some cases that, Neo-adjuvant administration has limited relapse-free survival benefit[16]. Thus, we have hypothesized that tumor subtype showing positive expression of TLR9 can induce inflammation response to released DNA motif from NACT derived damaged cells, and better survival response. In the present study, the prognostic importance of the expression of TLR 9 in tumor tissue treated with NACT has been investigated.

## 2 Material and Methods

### 2.1. Patients and samples

Samples of 42 unilateral primary breast carcinomas excised from women managed at Burdwan Medical College and Hospital were analyzed. The patients were diagnosed between 2016 and 2018. All 42 women underwent modified radical mastectomy, and 19 of them received neo-adjuvant chemotherapy (four cycles of FEC). None of the patients showed complete pathologic response (pCR) following NACT, and residual tumors were assessed in them. A qualified pathologist evaluated the histological subtype and the number of positive axillary lymph nodes. All the tumors were invasive carcinoma of no special type (NST). They were graded according to Scarff Bloom Richardson’s (SBR) grading system. To determine the prognostic value of TLR9 for breast cancer patients, two cohorts with NACT (n = 19) and without NACT (n = 23) were studied. Written consent was taken from all subjects included in the study. The study was approved by the institutional ethics committee, The University of Burdwan.

### 2.2. Immunohistochemistiy (IHC)

Tissue Sections of 4μm thickness were prepared from the paraffin-embedded tissue blocks. Tissue sections were deparaffinized in xylene and then rehydrated through a series of graded ethyl alcohol (100%, 90%, 80%, 70%, 50%, and 30%). Tissue sections were treated in citrate buffer (pH-6.8) for 15 min to retrieve the antigen in a microwave. After cooling, tissues were washed in distilled water, followed by 1X PBS [pH 7.4]. The endogenous peroxide activity was blocked with 3% H2O2 for 15 minutes. The samples were rinsed in 1X PBS and then pre-incubated with a protein blocking solution (10% Normal Goat Serum) for 1 hour in humidified chamber at 370 C. Human TLR 9 primary antibody (Thermo Fischer Scientific, Catalog – PA5–20202), was incubated (1: 100 dilution) at 4°C overnight in a humid chamber. Slides were washed three times in 1X PBST and incubated with HRP conjugated anti-rabbit secondary antibody (1:1000) for 30 min at room temperature. The peroxidase signal was visualized by treatment with a DAB substrate-chromogen system (Sigma, Cat No.- D3939) for 10 min. Finally, the sections were counterstained with Harrish-hematoxylin.

### 2.3. Evaluation of IHC staining for the expression of TLR9

TRL9 immunostaining slides were anonymized and blinded for the interpretation of the immunostaining score. A semi-quantitative intensity score (H score) modified from Measure et al.,(2015) [17] was used for TLR9 expression in malignant epithelial cells, briefly; score 0: negative staining, score 1: weak cytoplasmic staining, score 2: moderate cytoplasmic staining, score 3: strong cytoplasmic staining in most of the tumor cells, score 4: strong cytoplasmic and submembranous staining in almost all tumor cells. TLR9 underexpression was defined with an H score between 0 and 1 and TLR9 over-expression with an H score between 2 and 4. A similar semi-quantitative intensity score (H score) was developed for the expression of TLR9 in stromal cancer-associated fibroblast (CAFs) and mononuclear inflammatory cells (MICs) (score 0: negative staining, score 1: weak staining, score 2: strong staining). TLR9 under-expression was defined with an H score between 0 and 1 and TLR9 overexpression with an H score as 2.

### 2.4. Data mining through Publicly available datasets

To validate our IHC findings in other breast cancer cohorts, we have analysed different data sets from different studies available in different databases. The gene expression datasets were taken from Gene Expression Omnibus database [18, 19] and was used for prognosis and survival analysis.

#### 2.4.1. Validation as a marker, risk assessment and survivability analysis of using SurvExpress

SurvExpress (http://bioinformatica.mty.itesm.mx:8080/Biomatec/SurvivaX.jsp) is a web-based tool for survival anlysis and cancer-wide genome expression database with clinical outcome [20]. This platform uses TCGA and GEO database to draw survival plots for specific cancer type in relation to biomarkers. In performing survival and risk factor analysis of TLR9, we used data from the GEO dataset of GSE20685. This dataset originally contain data from 327 breast cancer patients. In this study, the gene expression profile was done using Affimatrix U133 plus 2.0 arrays [21]. The criteria of choosing the patient cohort and risk assessment is that all the patients’ cohorts must include survival data for over 30 samples, censoring indicators time to death, recurrence, relapse and metastasis. Statistical analysis was performed to compare survivability analysis in two defined risk groups. The high-risk and low-risk groups has been determined by splitting ordered prognostic index (PI) providing that each risk group have equal number of samples. The prognostic index of each sample was estimated using Cox-Survival analysis. A total of 164 samples have higher prognostic index than other 163 samples classified as high-risk group the other help is classified as low risk group. In our studied dataset, high risk group express significantly high level of TLR9 in comparison to low risk group. Cox proportional-hazards regression model was used to analyze the Kaplan-Meier log-rank analysis test and risk assessment.

#### 2.4.2. Survivability analysis of using Kaplan-Meier plotter

Kaplan-Meier plotter (http://kmplot.com/analysis/index.php?p=service&cancer=breast) is another web-based tool which analyses impact of 54,675 genes from 10,461 cancer samples using HGU133 Plus 2.0 array[22]. The main objective of this tool is to perform a meta-analysis-based biomarker assessment We mined previously published microarray data in the tool for 22,277 genes on breast cancer prognosis [23]. In this study, the correlation of the expression of TLR9 with neoadjuvant chemotherapy and survivability has been studied. Among the total cohort of 1402 patients (Affymetrix ID 223903_at), excluding the systemically untreated patients and patients receiving endocrine therapy, finally, 107 patients (GSE 16446) with neoadjuvant chemotherapy were selected for survivability analysis.

### 2.5. Statistical analyses

All statistical analyses were performed with Graph Pad Prism software package (Version 7.0). The correlation of TLR9 protein expression and different clinicopathological parameters and comparison between the expressions of TLR9 in the different patient cohorts was determined using the 2 x 2 contingency chi-square Fisher’s Exact Test. Cox proportional-hazards regression model was used to analyze the Kaplan-Meier log-rank analysis test and risk assessment In all tests, p-value < 0.05 was considered statistically significant.

## 3 Results

### 3.1. Differential TLR9 expression in breast tumors tissue based on IHC analysis

We first analyzed the expression of TLR9 in different grades of breast tumors. IHC results indicate that the TLR9 protein level was higher in both malignant epithelial cancer cells (Figure 1) as well as in adjacent stromal cells, which are mainly comprised of CAFs and MICs (Figure 2). We observed that TLR9 expression in malignant epithelial cancer cells was predominantly diffuse cytoplasmic with various intensity and sub-membranous (endolysosomal localization) localization in some cases in the tumors assigned with score 4 while the expression of TLR9 is confined to cytoplasmic areas in adjacent stroma. IHC results indicatedthat the expression of TLR9 protein is higher in the malignant epithelial cell (54.76%) than in the adjacent stromal compartment (11.9%).

**Fig. 1.**
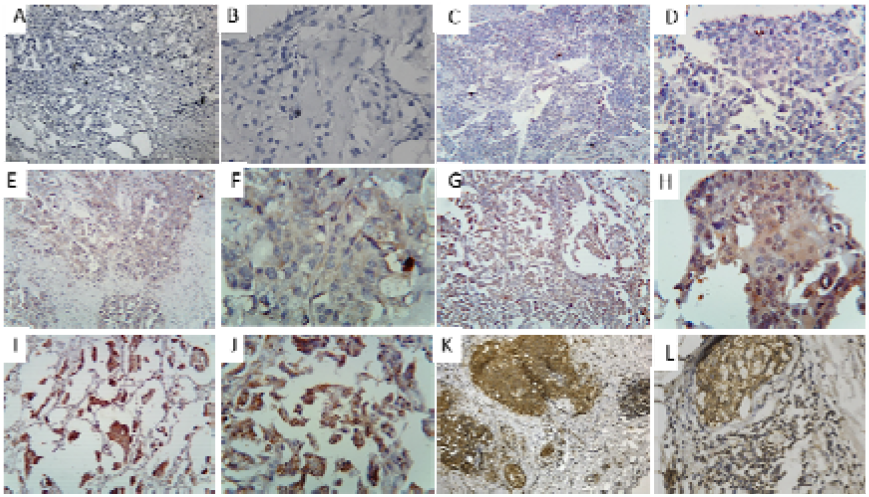
Representative micrographs depicting various scores of TLR9 expression in malignant epithelial cells. A) and B) Experimental Negative Control at 10X and 40X magnification, C) and D) IHC score 0 at 10X and 40X magnification, E) and F) IHC score 1 at 10X and 40X magnification G) and H) IHC score 2 at 10X and 40X magnification, I) and J) IHC score 3 at 10X and 40X magnification, K) and L) IHC score 4 at 10X and 40X magnification.

**Fig. 2.**
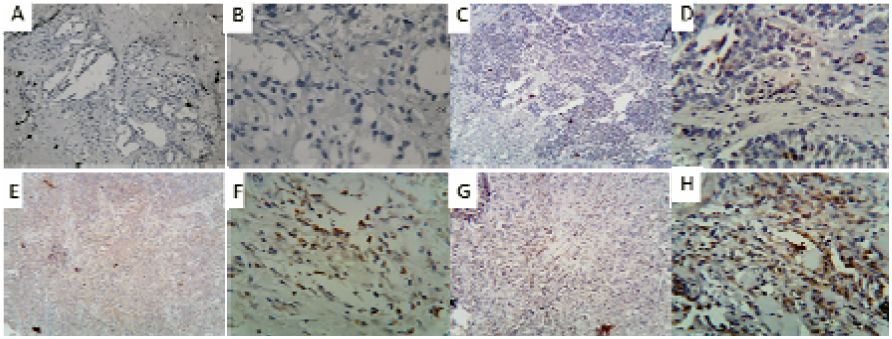
Representative micrographs depicting various scores of TLR9 expression in adjacent stroma of breast tumor tissue. A) and B) Experimental Negative Control at 10X and 40X magnification, C) and D) IHC score 0 at 10X and 40X magnification, E) and F) IHC score 1 at 10X and 40X magnification G) and H) IHC score 2 at 10X and 40X magnification.

### 3.2. Correlation of TLR9 expression with different clinicopathological parameters based on IHC analysis in breast tumor tissues

We analyzed the clinical significance of TLR9 protein expression in breast tumor tissue with several clinical parameters in malignant epithelial cell and adjacent stromal compartment; however, TLR9 expression was not found to be significantly correlated with any of the assessed clinicopathological parameters, such as age (p>0.99 and p = 0.64 respectively), histological grades (p = 0.76 and p = 0.95 respectively)and lymph node (p = 0.21 and p = 0.63 respectively) (Table 1).

**Table 1:** The association between TLR9 expression and clinical characteristics in breast cancer patients.

| Characteristics | TLR9<br>Positive<br>Score - ≥ 2 | TLR9<br>Negative<br>Score 0-1 | p-value |
| --- | --- | --- | --- |
| Malignant epithelial cells |  |  |  |
| Age |  |  |  |
| ≤ 50 | 15 | 13 | p>0.99 |
| ≥ 50 | 8 | 6 |  |
| Histological<br>grade |  |  | p=0.76 |
| I | 6 | 4 |  |
| II | 13 | 10 |  |
| III | 4 | 5 |  |
| Lymph node<br>status |  |  | p=0.21 |
| -ve | 11 | 9 |  |
| +ve | 12 | 10 |  |
| Adjacent stroma |  |  |  |
| Age |  |  |  |
| ≤ 50 | 4 | 24 | p=0.64 |
| ≥ 50 | 1 | 13 |  |
| Histological<br>grade |  |  | p=0.95 |
| I | 1 | 8 |  |
| II | 2 | 22 |  |
| III | 1 | 8 |  |
| Lymph node<br>status |  |  | p=0.63 |
| -ve | 4 | 16 |  |
| +ve | 1 | 21 |  |

### 3.3. Correlation of TLR9 expression in malignant epithelial cells with NACT based on IHC analysis in breast tumor tissues

Prognostic effect of expression of TLR9 in malignant epithelial cells was not studied in patients treated with NACT [24]. To assess the effect of NACT on expression of TLR9 and its prognostic effect, we compared the expression of TLR9 in patients treated with NACT and patients without NACT; we observed that the level of TLR9 in malignant epithelial cells was significantly high in patients treated with NACT compared to the patients without NACT (p = 0.0007) (Table 2).

**Table 2.** Expression of TLR9 in malignant epithelial cells of the patients without or with neo adjuvant therapy

| Expression of TLR9 in malignant epithelial cells | Patients without neo-adjuvant therapy | Patients with neo-adjuvant therapy | p-value |
| --- | --- | --- | --- |
| Negative (0-1) | 16 | 3 | 0.0007 |
| Positive (2-4) | 7 | 16 |  |

### 3.4. TLR9 expression in the stromal compartment was significantly associated with NACT based on IHC analysis in breast tumor tissues

Previously the prognostic significance of TLR9 expression in the stromal compartment of breast tumor has been reported (Sandholm et al., 2014). But the effect was not studied in patients treated with neoadjuvant chemotherapy. To assess the effect of neoadjuvant on the expression of TLR9 in the adjacent stroma and its prognostic effect, we compared the expression of TLR9 in patients treated with neoadjuvant chemotherapy and patients without neoadjuvant chemotherapy; however, no significant difference was found (p = 0.158) (Table-3a). On the other hand, we tried to establish a relation between expressions of TLR9 with the type of stroma. To assess the effect of neoadjuvant chemotherapy on type of stroma, we identified two types of stroma, the first type is desmoplastic that is hypercellular associated with numerous fibroblast and MICs and the second type is oedomatous or sclerotic that is hypocellular associated with low MICs infiltrate (Fig-3). We observed that increased in the expression of TLR9 in the stromal compartment increases the MIC infiltration and change the stromal condition in the patients given neoadjuvant chemotherapy (P = 0.0379) (Table-3b).

**Table 3a.** Expression of TLR9 in tumorstromal compartment of the patients without or with neo adjuvant therapy

| Expression of TLR9 in stromal compartment | Patients without neo-adjuvant therapy | Patients with neo-adjuvant therapy | p-value |
| --- | --- | --- | --- |
| Negative (0-1) | 22 | 15 | 0.158 |
| Positive (2) | 1 | 4 |  |

**Table 3b.**
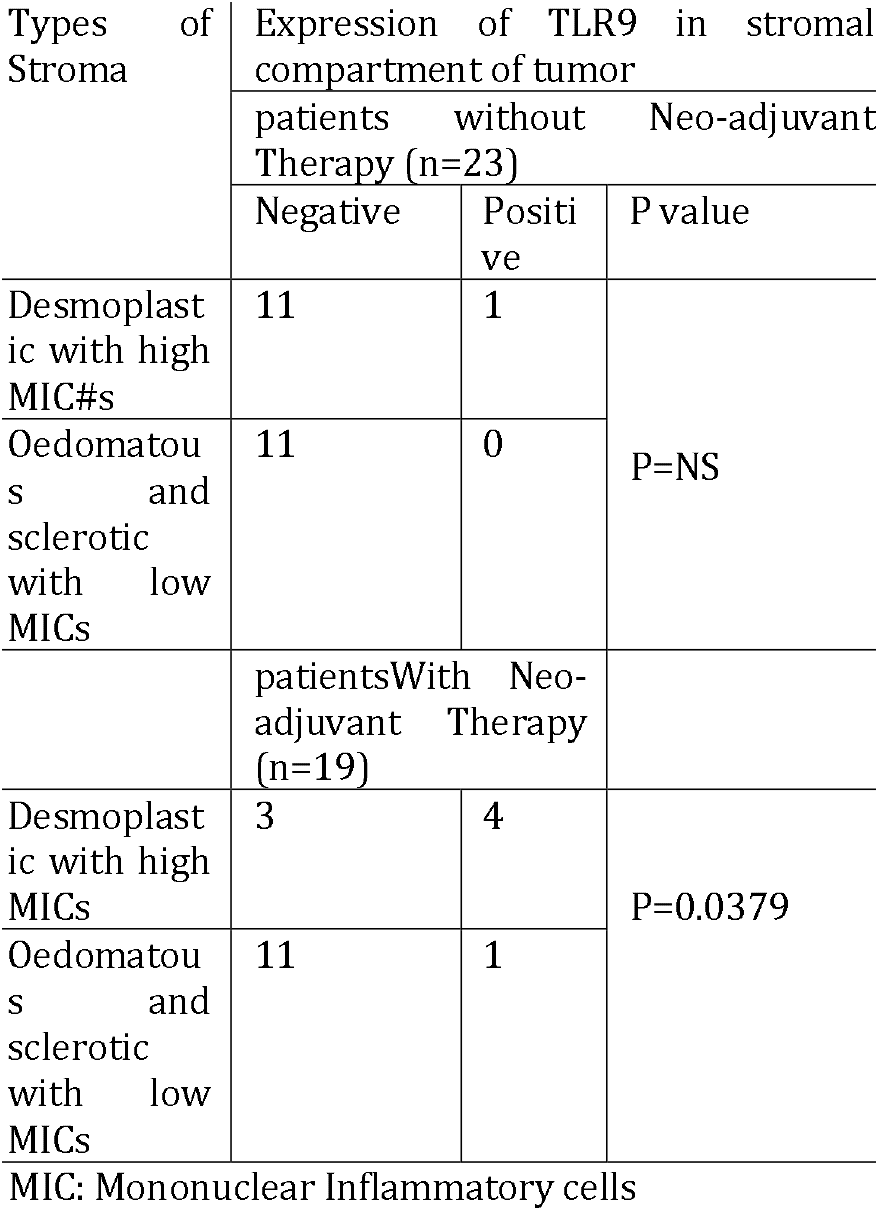
Expression of TLR9 in stromal compartment of tumor in relation with type of stroma of the patients without or with Neo-adjuvant Therapy

| Types of Stroma | Expression of TLR9 in stromal compartment of tumor |  |  |
| --- | --- | --- | --- |
|  | patients without Neo-adjuvant Therapy (n=23) |  | P value |
|  | Negative | Positive |  |
| Desmoplastic with high MIC#s | 11 | 1 | P=NS |
| Oedematous and sclerotic with low MICs | 11 | 0 |  |
|  | patientsWith Neo-adjuvant Therapy (n=19) |  |  |
| Desmoplastic with high MICs | 3 | 4 | P=0.0379 |
| Oedematous and sclerotic with low MICs | 11 | 1 |  |

MIC: Mononuclear Inflammatory cells

**Fig. 3.**
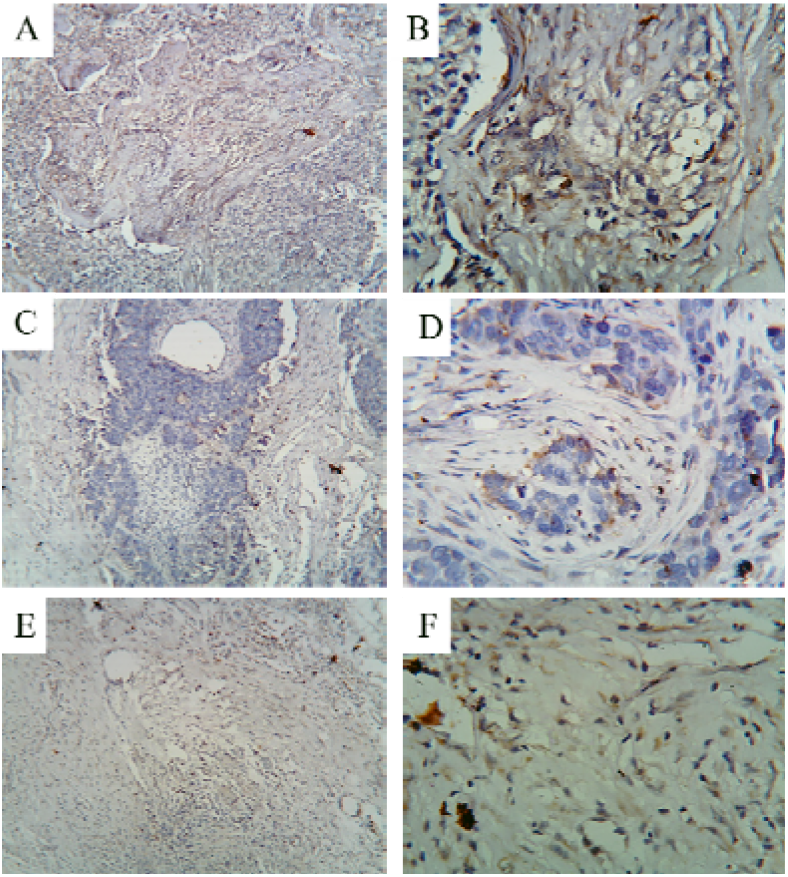
Immunohistochemical analysis of types of stroma. A) and B) Desmoplastic s troma at 10X and 40X magnification, C) and D) Oedomatous stroma at 10X and 40X magnification, E) and F) Sclerotic stroma at 10X and 40X magnification.

### 3.5. Validation of prognosis value of TLR9 and survivability with or without neoadjuvant chemotherapy

To determine relationship between the expression of TLR9 and clinical prognosis, we used web-based tools mainlyKaplan-Meier plotter and SurvExpress. We have analysed the survivability and prognostic value between patients treated with neoadjuvant chemotherapy and without neoadjuvantchemotherapy. Accordingly, it had been observed that there was no change in survivalship among the TLR 9 high or TLR 9 low expression group for patient cohort without neoadjuvant therapy in dataset of GSE 20685 (Figure 4A) analysed through SurvExpress. When we have taken the sample cohort of neoadjuvant therapy, there was a significant survivability benefit among the patient with higher TLR9 expression as analysed through Kaplan-Meier plotter for GSE 16446 dataset (Figure 4B). In case of risk assessment, it had been observed that, high risk group expressed, significantly high level of TLR9 in comparison to low risk group, but without neoadjuvant therapy, no survival benefit (Figure 4A).

**Fig. 4.**
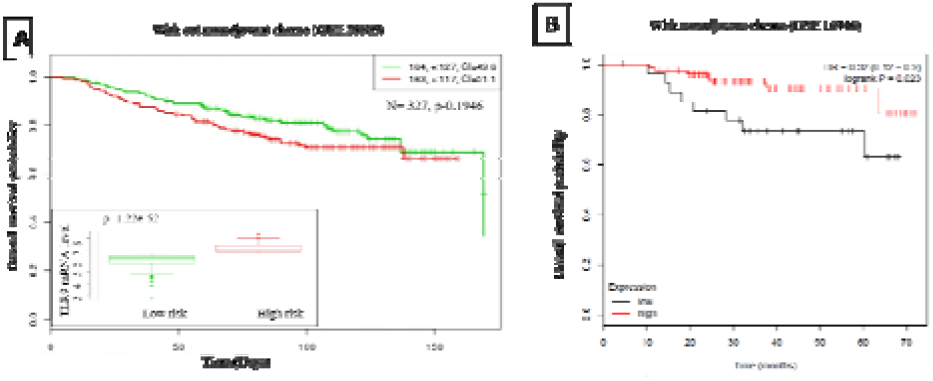
Expression of TLR9 and survivability in relation with or without neoadjuvant or chemotherapy. A) Kaplan-Meier analysis through SurvExpress database of overall survival for TLR9 expression under the condition without chemotherapy using breast cancer dataset GSE 20685. HR denotes hazard ratio. Insert, boxplot for the mRNA levels of TLR9 in high (red) and low (green) risk cohorts. A Cox p-value threshold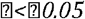 denotes level of significant. Risk assessment and expression of TLR9; high risk group expressed significantly higher level of TLR9 in comparison to low risk group (Inset). B) Kaplan-Meier survival analysis of the patients treated with neoadjuvant chemotherapy using microarray gene expression data (Data set GSE 16446) analyzed by KM plotter. High TLR9 expression has high overall survival in patients treated with neoadjuvant chemotherapy (Log Rank p = 0.023).

## 4 Discussion

In the present study, using IHC, expression analysis of TLR 9 in our indigenous sample cohort was studied. It was further validated by studying the prognostic importance of TLR 9 in a bigger sample size through using web-tools like SurvExpress and K-M Plotter.

TLR9 expression has been reported in various human tumors along with its correlation with several clinicopathological parameters of the cancer patients [10,11]. TLR9 has been reported to be over-expressed in different cancer both in-vitro as well as in human cancer samples, include breast cancer, neuroblastoma, gastric cancer, colorectal cancer, hepatocellular carcinoma, prostate cancer, and cervical squamous cell carcinoma [11]. In the present study, we analysed the significance of TLR9 expression in breast carcinoma tissues. TLR9 was expressed both in malignant epithelial cells as well as in adjacent stromal compartment and the expression of TLR9 in patients treated with neoadjuvant chemotherapy is significantly high. The dual localization of TLR9 wasendosomal as well as sub-membranous. In the present study, it has been observed that stroma becomes desmoplastic with high MIC infiltration at the site where there is a strong expression of TLR9 that has occurred within the tumor. In the clinical set-up, it has been shown that high tumor-infiltrating lymphocytes (TILs) at the time of diagnosis have a better response towards adjuvant trastuzumab. Breast tumorshave been reportedhaving weak immunogenicity, and the absence of TLR ligands have been shown to reduce the effectiveness of immune cells mediated adjuvant drugs [26].

However, the role of TLR9 in cancer progression in relation to Neoadjuvant chemotherapy has not been analyzed before. Neoadjuvant chemotherapy is used basically to downstage tumors and to eliminate the residual cancer burden in some patients. The residual cancer burden (RCB) defines the success of NACT, and it has been shown that residual cancer burden index (RCBI) or no residual disease (RCN) shows improved survival. NACT is given before any locoregional treatment like surgery or irradiation. NACT is mainly used during breast conservative surgery (BCS). Initially, NACT was used only in locally advanced breast cancer; however, it is now common for operable cancers also. A 9-year follow-up study ABCSG-07 trial showed shorter recurrence-free survival (RFS) between patients treated with NACT and patients treated with postoperative chemotherapy [27]. Data from randomly rolled trials (RCT) and other studies reported that the NACT administration has a similar OS [16] and DFS compared with that in adjuvant trials [28].However, Combined intratumoral administration of poly(I:C) and CpG in mice model of breast cancer trigger local inflammatory conditions that enhance the activity of systemic anti-ErbB2 mAb [26]. A phase II immunotherapeutic maintenance treatment in small cell lung cancer using TLR9 agonists Lefitolimod have positive overall survivability [2 9]. Another phase I clinical trial using lefitolimod in combination with CTLA-4 checkpoint inhibitor ipilimumab in advanced malignancies is under progress [30]. The possible reason for using TLR9 agonists in cancer therapy was based on their ability to activate the inflammatory response to activate the immune system. The inflammation may amplify antitumor immune response, eradication of tumors, and translate into a better prognosis in breast cancer [10].

To improve our understanding of the value of TLR9 as prognostic marker in breast cancer patients in association with neoadjuvant chemotherapy, we next investigated the association between the expression level of TLR9 and survival in various breast cancer datasets. The prognostic value ofTLR9 mRNA expression was assessed using Kaplan-Meier plotter and SurvExpress. In SurvExpress analysis, high expression of TLR9 was found in the high-risk patients compared with those in low-risk patients. High expression of TLR9 expression does not predict any difference in survival in patients not treated with neoadjuvant chemotherapy. Further to justify our hypothesis, the survival of the patients was studied using the Kaplan-Meier Plotter using the GEO data set (Affymetrix ID 223903_at TLR9). It was revealed with KM analysis that the overall survival of the patients has no correlation with TLR9 expression in overall condition, but inpatients treated with neoadjuvant chemotherapy, survival increases with an increase in TLR9 expression which was studied using dataset GSE 16446. Immuno-sensitization of the tumor using TLR9 ligand in neo-adjuvant set-up leads to an increase in infiltrating immune cells that might help the other drugs. This might be due to DNA fragments derived from chemotherapy-treated dead cancer cells that may be taken up by the surviving cancer cells. These DNA fragments can induce TLR9 as damage-associated molecular pattern (DAMP) ligand that may contribute to treatment responses via tumor TLR9 expression with augmenting anticancer immune response.

In conclusion, we have shown that neoadjuvant chemotherapy induces high TLR9 expression in IBCs. High TLR9 expression is correlated with high immune cell infiltration in the tumor and produces an inflammatory microenvironment that may enhance the activity of other therapeutic agents and might also help in the establishment of longer-lasting antitumor immunity. TLR9 expression is also good for the survival benefit of the patients treated with neoadjuvant chemotherapy. Thus, high TLR9 expression may act as a good prognostic marker in the patients receiving neoadjuvant chemotherapy.

## Data Availability

Relevant data will be provided as per request basis

## Acknowledgments

The authors acknowledge Dr. Sourav Sau, MD, DNB, Department of Radiotherapy, Burdwan Medical College, and Hospital for providing the basics and data related to the neo-adjuvant chemotherapy of the patients. The authors acknowledge the Principal, Burdwan Medical College, and Hospital for the support to carry out this study. The authors also acknowledge Mr. KartickKarmakar, Medical Technologists, Department of Pathology, Burdwan Medical College, and Hospital for his technical support in this study.

